# Immune response against SARS-CoV-2 of primary healthcare personnel in a commune of Santiago, Chile: follow-up at 6 months

**DOI:** 10.1101/2022.08.03.22278369

**Authors:** A. Olea, I. Matute, M. Hirmas, C. González, M. Iruretagoyena, J. Munita, E. Pedroni, MI. Gómez, L. Cortés, J. Hormazábal

## Abstract

**Background:** The COVID-19 pandemic that emerged in Wuhan, China at the end of 2019, spread rapidly around the world with almost 600 million cases and 6.3 million deaths today. The most affected were health workers with at least three times the risk of contracting the disease than the general community. Most studies on seroprevalence in health workers focus on hospital care establishments and what happens in Primary Health Care (PHC) has not been investigated with the same intensity.

**Objectives:** to determine the prevalence and know the variation of antibody titers for SARS-CoV-2 in serial samples of primary healthcare personnel from the commune of La Pintana.

**Method:** an analytical observational study with a cross-sectional and a longitudinal component, carried out from November 2020 to June 2021. The first component consisted of an IgG antibody seroprevalence study performed at baseline (time 0) in volunteer of a universe of 900 workers. The longitudinal component considered the monitoring of IgG antibodies in those who presented a positive result at baseline and the analysis of neutralizing antibodies in a random sub-sample of 50% of them. Additionally, sociodemographic and clinical information was collected via a questionnaire. Univariate, bivariate, and longitudinal analyses were performed to evaluate differences in antibodies. The study was approved by the Universidad del Desarrollo’s Scientific Ethics Committee.

**Results:** 463 primary healthcare workers participated, mostly women and with a median of 38 years; doctors and nurses represented 9.5% each and 14.7% had a history of COVID-19. The seroprevalence at baseline was 22.3% and was associated with younger age, being a doctor and having been in close contact of a case. IgG titers increased with the vaccine, but decreased over time. At the 6-month follow-up, 76% had neutralizing antibodies. Those belonging to indigenous peoples had higher IgG levels and higher rates of neutralizing antibodies.

**Conclusion:** Healthcare workers were highly affected by COVID-19, and the medical profession and younger age were factors associated with increased risk. Antibodies decrease over time, highlighting the importance of follow-up studies, as well as the importance of vaccination boosters in healthcare workers, especially those in PHC.

**Financing:** This project was funded by the Universidad del Desarrollo, COVID19-UDD 2020-21 Funds.

## Introduction

In December 2019, an outbreak of serious respiratory infections caused by a previously unknown strain of coronavirus, which was called SARS-CoV-2 (1) and the disease it causes, COVID-19 (Coronavirus disease-19), was reported in China (Wuhan). Cases spread rapidly, and on March 11, 2020, the World Health Organization (WHO) declared it a pandemic. Two years after that milestone, there are more than 565 million cases and 6.4 million deaths worldwide, with more than 11.8 billion vaccines administered. (2) In Chile as of July 2022, there are more than 4 million cases and almost 60,000 deaths. (3) The vaccination campaign against SARS CoV-2 began in January 2021, reaching full vaccination coverage (including boosters) of 92% of the target population by July 2022. (4)

The transmission characteristics of the disease implied a great demand for healthcare workers worldwide, who have needed to perform in the first line of care, thus being exposed to a greater risk of disease. (5) Health personnel represent less than 3% of the total population; however, between 14% and 35% of reported COVID-19 cases were healthcare workers. (6) The World Health Organization estimates that around 115,000 health workers have died worldwide. (7) Therefore, in August 2020, the Pan American Health Organization (PAHO) urged countries to strengthen health service capacities and keep personnel equipped with resources and training to deliver an adequate and timely response to the pandemic. (8)

There is abundant evidence on seroprevalence in health professionals; however, this is mainly focused on hospital care facilities. (9–11) Research on primary care personnel is scarce, although it has increased in the last year. (11–13) Internationally, these studies report seroprevalence close to 5% during the first wave and between 8.1% and 17% during the second, figures significantly higher than in the general population. (14,15)

In a study conducted on health workers at a large hospital in Sweden, the seroprevalence of IgG antibodies against SARS-CoV-2 was 19.1%. (16)

In Chile, a nationwide seroprevalence study, which included more than a third of public sector healthcare personnel, both at the hospital and primary care levels, reported total seropositivity of 7.2% after the first wave; hospital-level workers had twice the risk compared to those in primary care (seroprevalence of 9.4% and 4.4%, respectively). (17) Another study in long-term care facilities for the elderly in three regions of Chile recorded 6.3% seropositivity in health officials, showing a positive and significant correlation between the seroprevalence of staff and residents. (18) In a study conducted in a hospital in Santiago, Chile, the seroprevalence of those who worked in the COVID patient area was 24% (43% asymptomatic).(19)

The IgG-type humoral immune response appears between 15 and 21 days from the time of infection (20) and neutralizing antibodies (NAbs) correspond to a portion of IgG antibodies that have the ability to neutralize the virus, preventing its entry into cells. They are a vital part of the immune response, and their measurement is fundamental to know the immunity produced either by infection or by vaccine. (21)

The detection of antibodies presents a sensitivity close to 100% in the convalescence and subsequent phases (22,23). International studies show that anti-SARS-COV-2 IgG antibody titers are detectable at high levels for up to 3 months, decreasing between 6 and 9 months. (24–26) Other studies demonstrated a persistence of IgG and neutralizing antibodies for up to 13 months, the concentrations of which remained higher in people with significant symptoms compared to people who were asymptomatic or with mild upper respiratory tract discomfort. (26) A study on the persistence of neutralizing antibodies (NAbs) among healthcare workers showed a significant decrease in NAbs titers between day 21 of infection and the third month (p < 0.0001). (27) In a post-vaccine follow-up study, neutralizing antibodies persisted for 6 months after the second dose, as detected by three different serological assays. (28)

The objective of our study was to know the variation of antibody titers for SARS-CoV-2 in serial samples of primary healthcare personnel from the La Pintana commune, through the measurement of IgG antibodies and neutralizing antibodies (NAbs) in a subsample.

## Method

### Study design, population, and sample

An analytical observational study was conducted with a cross-sectional and a longitudinal component. The target population corresponded to the total number of workers (N=900) who worked from November 2020 to June 2021 in the six primary healthcare (PHC) facilities in the La Pintana commune, a sector of high social vulnerability located in the south of the Chilean capital and which presented high incidence rates of cases and deaths during the first wave. (3)

The cross-sectional component consisted of a seroprevalence study of antibodies against SARS-CoV-2, conducted at the beginning of the study (time 0), for which a minimum sample of 381 people was estimated, with 95% confidence, 3% error, and an IgG+ seroprevalence of 19%; (16) volunteer sampling was used, and all staff members were invited to participate. The longitudinal component included those who presented an IgG+ result at time 0. A subsample of 50% of them was taken randomly for neutralizing antibody analysis at the end of the study (time 3).

### Variables and data collection

For the determination of the serology of antibodies against SARS-CoV-2, venous blood samples were collected. The serum was separated, divided into aliquots, and stored at −20°C until analysis.

Specific IgG was determined using commercial enzyme-linked immunosorbent assays (ELISA), COVID-19 ELISA IgG (Vircell, Grenada, Spain) using recombinant SARS-CoV-2 nucleocapsid (N) protein and spike glycoprotein (S). Previously, the trial was validated with serum samples from 60 COVID-19 patients confirmed by RT-PCR and 40 asymptomatic RT-PCR negative patients. The Vircell trial had a sensitivity of 85.7% and a specificity of 98.5% after 3 weeks of symptom onset. The test was performed according to the manufacturer’s instructions. (29)

The cut-off point for IgG+ was >=8. We also included two participants with a limit IgG result but with a recent medical history of COVID-19 confirmed with a positive RT-PCR test.

The activity of the SARS-CoV-2 specific neutralizing antibodies in serum samples was determined by neutralization assays, using replication-competent pseudotyped viral particles. (30) Vero E6 cells were seeded as a monolayer in a 96-well optical bottom plate and incubated overnight at 37°C with 5% CO2. On the following day, each serum was two-fold serially diluted and incubated with pseudovirus VSV-GFP-Spike SARS-CoV-2, at 37°C for 30 min. Then, the virus-serum mixture was transferred to a 96-well plate containing the Vero cells and incubated at 37°C with 5% CO2 for 18-20 h. After incubation, the cells were washed and fixed with paraformaldehyde 4%. The infection was measured as GFP fluorescence intensity using a Cytation3 plate reader. The half-maximal inhibitory concentration (IC50) of the NAbs was calculated using nonlinear regression analysis in the Graphpad software, with each sample tested in triplicate.

The serology and neutralizing antibody analyses were carried out in the laboratory of the Institute of Sciences and Innovation in Medicine (ICIM) in the Faculty of Medicine of the Universidad del Desarrollo-Clínica Alemana.

In addition, each participant underwent face-to-face interviews using structured questionnaires, designed to collect sociodemographic variables: sex, age, belonging to an indigenous people, nationality (Chilean or other), health system (public or private) and occupation (medical or other), as well as health: having suffered an episode of illness, history of COVID-19 with PCR+ and vaccination record. Study data were collected using Tablet and managed using REDCap electronic data capture tools hosted at Universidad del Desarrollo. (31,32) REDCap (Research Electronic Data Capture) is a secure, web-based software platform designed to support data capture for research studies, providing 1) an intuitive interface for validated data capture; 2) audit trails for tracking data manipulation and export procedures; 3) automated export procedures for seamless data downloads to common statistical packages; and 4) procedures for data integration and interoperability with external sources.

The data collection was carried out four times. Time 0 (T0) corresponded to the measurement used for the cross-sectional study and was executed in November 2020. Then three more measurements were conducted for those people with a positive result at time 0: time 1 (T1) January 2021, time 2 (T2) March 2021 and time 3 (T3) June 2021. From this last measurement, 50% of the samples were selected for the analysis of neutralizing antibody levels.

Trained nurses conducted both the blood sample collection and the application of questionnaires, after the informed consent process.

### Data analysis

The SPSS v.27 statistical package was used for the analysis. First, a descriptive analysis of the participants, their risk factors and disease episodes were performed. For quantitative variables, measures of central tendency and dispersion were calculated, and for categorical variables, frequencies and percentages were determined. To study factors associated with the disease, a multivariate analysis was performed using logistic regression, considering a p<0.05.

Antibodies against SARS-CoV-2 were analyzed in each measurement, by counting IgG as a quantitative variable and from its categorization as positive or negative.

To compare IgG concentrations in T1 and T3, the Wilcoxon Test was used, and for the longitudinal analysis of IgG counts, a Generalized Linear Model (GLM) was developed, considering p<0.05.

For neutralizing antibodies, a descriptive analysis was carried out based on the quantitative count and the categorization as positive or negative.

### Ethical considerations

The study was approved by the Scientific Ethics Committee of the Universidad del Desarrollo. Participation was voluntary, the process of signing informed consent was conducted, indicating the scope of the study. There were no associated risks or costs for the participants, and as a benefit, the subjects obtained results on their levels of IgG antibodies against SARS-CoV-2. The result of the blood sample was reported in a maximum of 14 working days to each participant. Data privacy was safeguarded at all times using encrypted information that could only be accessed by researchers, under security standards with a personal and non-transferable key.

## Results

### Cross-sectional component

The cross-sectional component (T0) had the participation of 463 primary healthcare officials from the commune of La Pintana (51.4% of the total), with a median age of 38 years (range 18-83 years), a greater proportion of women (75.8%; 351/463) and beneficiaries of public health insurance (Fonasa) (66.5%; 306/460). Doctors and nurses represented 9.5% (44/463) each, 49.9% corresponded to other health professionals (231/463) and 31.1% (144/463) to personnel from other areas.

A 14.7% (68/463) had a history of COVID-19 by PCR, and the seroprevalence of antibodies against SARS-CoV-2 at T0 was 22.3% (103/463). Older age appears as a protective factor (OR 0.96; CI95% 0.94-0.98), while being a doctor (OR 3.33; CI95% 1.71-6.51) and have been in close contact (OR 2.17; CI95% 1.30-3.62), are risk factors against transmission.

Regarding the clinical picture, 22.3% were asymptomatic (23/103) and among those who had symptoms (77.7%), the most frequent were headache (66%; 68/103), myalgia (57.3%; 59/103) and odynophagia (46.6%; 48/103). The probability of positive serology was greater than 70% in those who had anosmia or ageusia, and only 5 people were hospitalized for COVID-19 (4.8%; 5/103).

The median antibody titers was 4.7; 4.0 for negative cases (RIQ 2.8-13.1) and 20.9 for positive cases (RIQ 13.3-33.9), indicating a statistically significant difference (p=0.029).

### Longitudinal Component

#### Participant Characteristics

Of the 103 positive cases, 100 completed follow-ups (97.1%). Of these, 79% (79/100) were women and the median age at the beginning of the study was 32 years (range 27-39 years) (Table 1).

**Table 1.** Sociodemographic and clinical characteristics of primary healthcare workers with igG+, La Pintana commune, Metropolitan Region, Chile 2020.

|  |  |
| --- | --- |
| **Total** | **100(100%)** |
| Women | 79(79%) |
| Men | 21(21%) |
| Age (median and IQR) | 32(27-39) |
| Indigenous | 13(13%) |
| Non-indigenous | 87(87%) |
| Chilean nationality | 87(87%) |
| Other nationality | 13(13%) |
| FONASA (Public healthcare system) | 61(61%) |
| ISAPRE (Private healthcare system) | 39(39%) |
| Physician | 20(20%) |
| Nurse | 14(14%) |
| Other profession or occupation | 66(66%) |
| Presented a respiratory infection episode | 77(77%) |
| Did not present a respiratory infection episode | 23(23%) |
| History of COVID-19 by PCR | 65(65%) |
| No history of COVID-19 by PCR | 35(35%) |
| Vaccinated with two doses at T3 | 88(88%) |
| Vaccinated with one dose at T3 | 3(3%) |
| Not vaccinated at T3 | 9(9%) |

At T0, none of the participants had begun the vaccination process; at T1, only one person had the first dose of the vaccine; at T2, 16% (16/100) had received a first dose and 69% (69/100) had already registered the second dose; at T3, the percentage with a second dose reached 88% (88/100), 3% had one dose (3/100) and 9% remained unvaccinated (9/100) (Table 1). As for the vaccine administered, 88 subjects received Sinovac (96.7% of those who had at least one dose).

#### Antibodies to SARS-CoV-2

In the first antibody measurement, the IgG count had a median of 20.55, which increased to 24.9 at time 1, without this variation being statistically significant. When comparing the IgG counts of time 1 and time 2, a significant increase was recorded (p<0.001), and then also significantly decreased (p<0.001) at time 3. The seroprevalence of antibodies against SARS-CoV-2 remained over 95% at all times (Table 2).

**Table 2.** Concentration of IgG in primary healthcare workers at each follow-up time and neutralizing antibodies measurement against the spike of SARS-CoV-2 at T3, La Pintana commune, Metropolitan Region, Chile 2020.

| Follow-up time | Mean | Dev. Deviation | Median | IQR | Minimum | Maximum | Range | N | Seroprevalence |
| --- | --- | --- | --- | --- | --- | --- | --- | --- | --- |
| T0 | 26.53 | 17.95 | 20.55 | 13.35-33.95 | 6.70* | 85.20 | 78.50 | 100 | 100% |
| T1 | 29.61 | 19.40 | 24.90 | 11.32-56.2 | 6.50 | 56.30 | 49.80 | 100 | 95% |
| T2 | 43.30 | 25.13 | 40.80 | 17.1-75 | 2.60 | 75.00 | 72.40 | 99 | 97% |
| T3 | 35.68 | 20.70 | 31.05 | 15.2-58.58 | 1.60 | 75.00 | 73.40 | 100 | 99% |
| NAbs in T3** | 1312.56 | 2574.32 | 460.58 | 29.08-1553.61 | 0.00 | 12632.64 | 12632.64 | 50 | 76% |

\* A participant with an IgG limit result but with a confirmed RT-PCR COVID-19 medical history was positive in the next sample collection, so this person was included in the analyses.

When comparing the results of time 3 (91% vaccinated) and time 1 (without vaccine), the median IgG increased, which was a statistically significant difference (p<0.001), and 5 people who had negative serology at T1 became positive at T3, while only one person remained negative.

The longitudinal analysis, considering the entire period, shows the commented fluctuations, with the time factor being statistically significant (p<0.001) (Figure 1).

**Figure 1.**
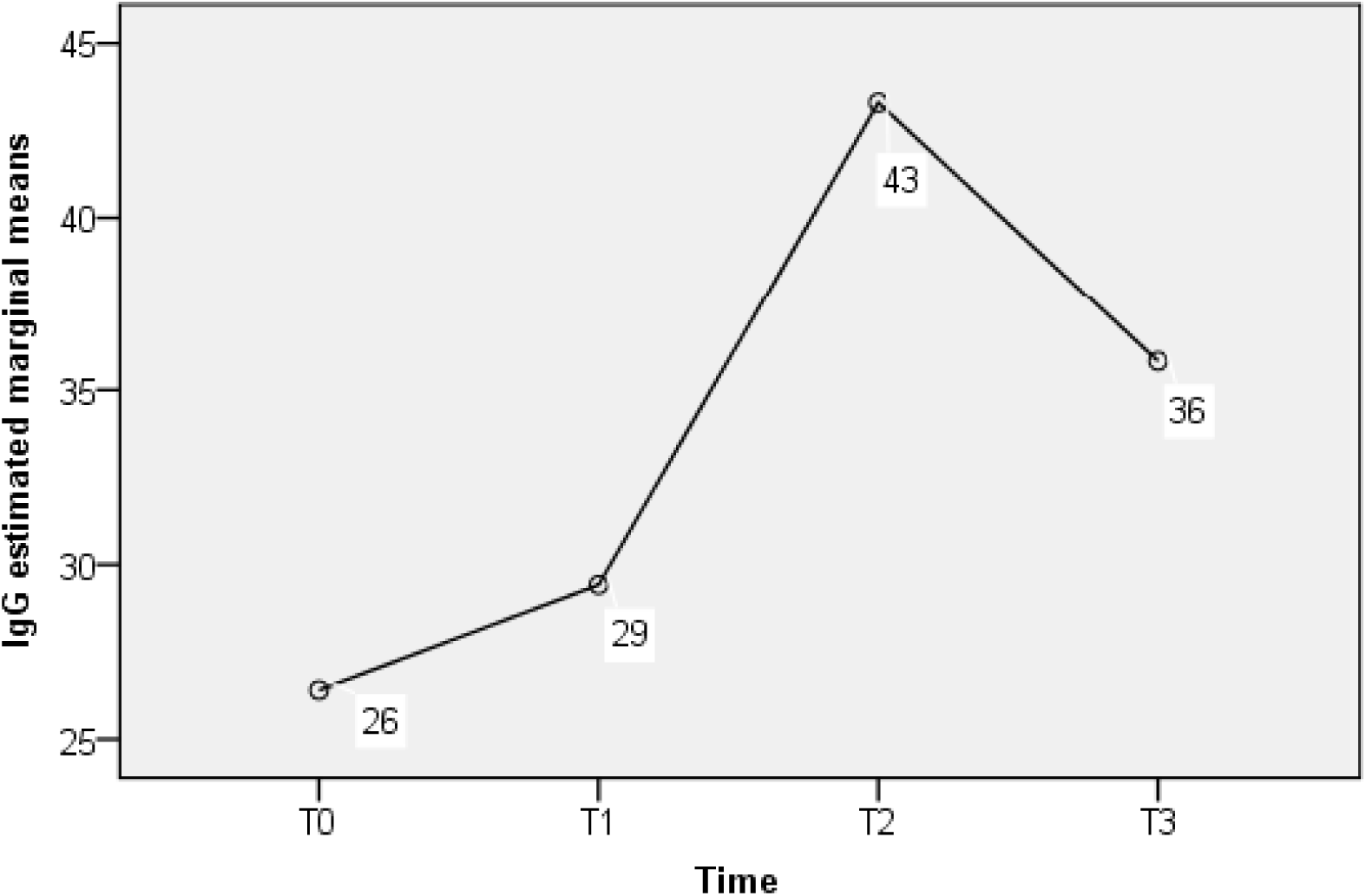
Concentration of IgG against SARS-CoV-2 in primary health care workers at eeach follow-up time. La Pintana commune, Metropolitan Region, Chile 2020.

When specifically analyzing the concentration of IgG at T3 according to other variables, significant differences were recorded (p=0.021) in favor of those who indicate belonging to an indigenous people (n=13, median=58.7, RIQ=20.65-58.95) compared to those who do not (n=87, median=26.4, RIQ=14.9-58.4) (Figure 2). Having had a symptomatic episode of COVID-19 did not present statistical significance, despite the fact that their median IgG was higher (32.5) than that of asymptomatic people (20.8). Regarding vaccination, there were also no significant differences between those who received the vaccine and those who did not.

**Figure 2.**
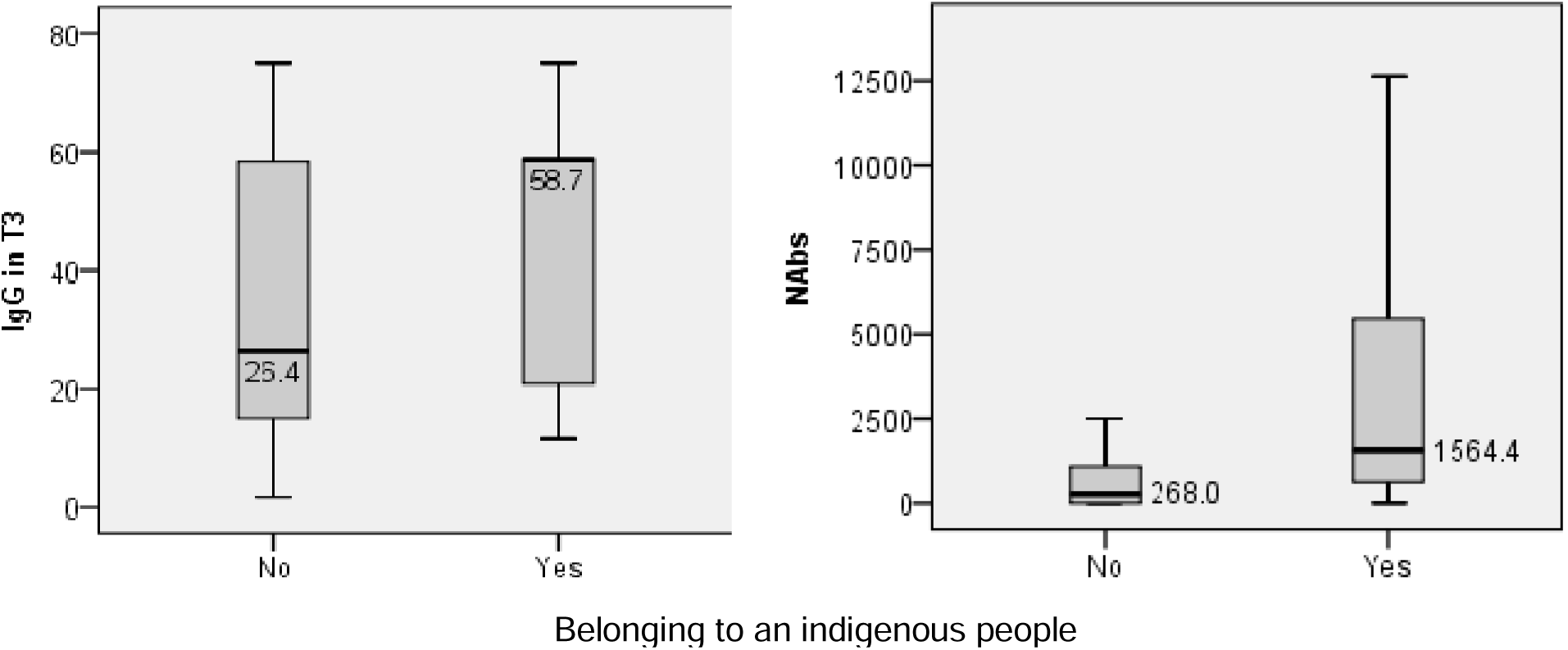
IgG and Nabs concentration against SARS-CoV-2 in primary healthcare workers at T3, according to belonging to an indigenous people, commune of La Pintana, Metropolitan Region, Chile 2020.

#### NAbs

As observed in Table 2, the NAbs count in the 50 aliquots analyzed at time 3 presented a median of 460.58 (IQR 29.08-1553.61), with a positive and significant correlation to the total antibodies against SARS-CoV-2 (p<0.001). The bivariate analysis of the count according to demographic and clinical variables only shows significant results in terms of ethnicity (p=0.027), with a higher median in people who belong to an indigenous people (n=5, median 1564.41, RIQ 566.1-6429.56) compared to those who do not (n=45, median 295.98, RIQ 0.0-1211.36) (Figure 2).

Again, no differences were observed in terms of symptomatology or vaccination status; however, the median was higher in people vaccinated with some dose (501.5), than in those three cases that remained unvaccinated (145.29).

Finally, when categorizing the presence of NAbs, 76% were classified as positive (38/50).

## Discussion

The prevalence found (22.3%) is very similar to that reported in a study conducted in healthcare workers at a hospital in Santiago from April to July 2020 (24%) (19) and consistent with previous evidence in health personnel, whose seroprevalence is higher than what is observed in the community, demonstrating the high risk faced by these personnel. (33–38)

When controlling for potential confounding variables in our study, the medical profession increases the chance of disease. This finding is probably associated with the greater time that these professionals spend in PHC providing care to people with recent onset of the disease, when there is a greater risk of transmission. (39)

Likewise, and consistent with the literature, younger age is a risk factor of disease, as well as having completed quarantine due exposure to a confirmed case. (36)

Regarding the symptomatology, only 30% of the cases that reported COVID-19, had a fever, which was a very non-specific symptom. On the other hand, a third of people with COVID-19 present ageusia or anosmia; the positive predictive value reaches 74%, which means that a person with one of these two symptoms had a high probability of being a disease case. This is relevant when establishing and adjusting case definitions.

During the follow-up, vaccination against SARS-CoV-2 was implemented, which did not allow us to know the natural evolution of the antibodies produced by the disease, but we obtained post-vaccination antibody titers. Only the sample taken in January 2021 (T1) compared to that of November 2020 (T0), allows seeing the natural variation of IgG concentrations and where 5% of people remained negative in the two months elapsed.

At all times of follow-up and consistent with that described in other studies, (40) those over 60 years of age, asymptomatic and those not belonging to an indigenous people, presented lower IgG titer levels. In several studies, antibody prevalence was higher in black and Asian health workers compared to white. (41–43) In addition, in our study, women had a lower IgG concentration during follow-up.

Similar to other international studies, (24–26) the highest concentration of IgG antibodies was obtained one month after vaccination (T2), decreasing at 3 months (T3), despite the fact that 88% had two doses of CoronaVac. This is consistent with the results of a previous study conducted in Chile, where IgG seropositivity decreased over time since vaccination for those who received CoronaVac and not for those who received BNT162B2 vaccine (Pfizer-Biontech). (44)

Regarding neutralizing antibodies (NAbs), these were present in 76% of people at 6 months of follow-up, with a higher concentration in those who had clinical disease (PCR+) compared to the group of asymptomatic individuals, a finding consistent with international literature. (22)

It is interesting to note that at the end of the follow-up, although IgG seroprevalence was 99%, only 76% had Nabs, a fact that could be due to what was previously commented about the CoronaVac vaccine.

Regarding the limitations of our study, it is first recognized that the sampling strategy was by invitation, and there may be self-selection of those at higher risk or who presented COVID symptoms and, therefore, were interested in knowing their immune status. Another limitation, common to survey-based studies, is the fact of answering what is “expected or desirable”, and risk behaviors may be underestimated. Finally, it asked about activities or behaviors from the 8 months prior to the study, which could introduce memory bias.

Conclusions: Healthcare workers were greatly affected by COVID-19, and the medical profession and younger age were factors associated with an increased risk. The concentration of antibodies was higher in those who belong to an indigenous people. Antibodies decrease over time, pointing to the importance of follow-up studies, as well as the importance of vaccination boosters for healthcare workers especially in PHC.

## Data Availability

All data produced in the present work are contained in the manuscript

## Acknowledgments

The authors thank the health workers participating in this study from the Commune of La Pintana; Paulina Reinoso Ríos, the head of the Municipal Health Department of La Pintana; and Gonzalo Cartagena, Melissa González and Constanza Vega, professionals from the Municipal Health Department of La Pintana.

Thanks also are due to Claudia Pérez, the head of the field team; Nicole Rodríguez and Catalina Ruz, the team of nurses in charge of gathering the information in the field; and Lina Rivas, the laboratory assistant. We also kindly thank Isidora Castillo for her collaboration in reviewing evidence for this manuscript.

